# Aberrant Activation of the Mentalizing Brain System During Eye Gaze Discrimination in Bipolar Disorder

**DOI:** 10.1101/2021.04.19.21255514

**Authors:** Ivy F. Tso, Cynthia Z. Burton, Carly A. Lasagna, Saige Rutherford, Beier Yao, Scott J. Peltier, Timothy D. Johnson, Melvin G. McInnis, Stephan F. Taylor

**Affiliations:** Department of Psychiatry, University of Michigan, Ann Arbor; Radboud University Medical Centre, Nijmegen, The Netherlands; Department of Psychology, Michigan State University; Functional MRI Laboratory, University of Michigan, Ann Arbor; Department of Biomedical Engineering, University of Michigan, Ann Arbor; Department of Biostatistics, University of Michigan, Ann Arbor

**Keywords:** bipolar disorder, social cognition, fMRI

## Abstract

Bipolar disorder (BD) is associated with a range of social cognitive deficits. This study investigated the functioning of the mentalizing brain system in BD probed by an eye gaze perception task during fMRI. Compared with healthy controls (*n* = 21), BD participants (*n* = 14) showed reduced preferential activation for self-directed gaze discrimination in the medial prefrontal cortex (mPFC) and temporo-parietal junction (TPJ), which was associated with poorer cognitive and social functioning. Aberrant functions of the mentalizing system should be further investigated as marker of social dysfunction and treatment targets.

**Highlights:**

- Social dysfunction in bipolar disorder (BD) may be due to altered mentalizing.
- Individuals with BD showed reduced activation in the mentalizing brain system.
- Aberrant activity of the mentalizing system was associated with poorer functioning.

## 1. INTRODUCTION

Bipolar disorder (BD) is a serious psychiatric condition that affects more than 45 million people worldwide and frequently leads to persistent social and occupational disability (James et al., 2018; Sanchez-Moreno et al., 2009). Deficits in social cognition, or “the mental operations that underlie social interactions, including perceiving, interpreting, and generating responses to the intentions, dispositions, and behaviors of others” (Green et al., 2008), significantly contribute to the significant functional impairment observed in BD (Vlad et al., 2018), and have emerged as a critical target to improve social functioning.

Large meta-analytic studies have shown that facial emotion processing and theory of mind/mentalizing (representing the ability of others to take a psychological perspective) are crucial components of social cognition consistently impaired in individuals with BD (Bora et al., 2016; Samamé et al., 2012). Relatedly, eye gaze processing is a particularly relevant ability as it involves processing gaze direction in the context of faces to make inferences about the attention and intention of others. Dysfunctional gaze perception has been observed in BD (Berchio et al., 2017) and may be due to a self-referential bias—the tendency to interpret the behavior of others in relation to the self (Yao et al., 2018). Altered gaze processing may be amplified for individuals with BD who experience psychosis, as there is some evidence to suggest that social cognitive deficits are more severe in psychotic BD (Thaler et al., 2013), though these processes have not been comprehensively investigated to date.

Limited prior work has examined the neural correlates of mentalizing among individuals with BD, with variable and inconclusive results, likely due to differences in the tasks used to probe mentalizing. The brain circuits supporting mentalizing have been extensively studied (Frith and Frith, 2006; Gallagher and Frith, 2003; Molenberghs et al., 2016; Van Overwalle and Baetens, 2009). While a number of brain regions have been implicated (e.g., precuneus, superior temporal sulcus, cingulate cortex, inferior frontal gyrus, temporal poles), the temporoparietal junction (TPJ) and medial prefrontal cortex (mPFC) have emerged as two critical regions (Molenberghs et al., 2016; Van Overwalle, 2009). In studies of mentalizing in BD, reduced activation in limited cortical regions using a theory-of-mind task featuring a geometric shape as active agents has been found (Malhi et al., 2008), while others reported higher levels of neural activation in key regions, possibly reflecting a compensatory response, on theory of mind tasks assessing self vs. other processing (Grant et al., 2018). Only one recent study has specifically examined TPJ and mPFC and found, using a cartoon-based task making inference of the characters’ affective mental state, reduced bilateral activation of the TPJ and as diminished coupling between the bilateral TPJs and the mPFC in individuals with euthymic BD as compared with healthy controls (Willert et al., 2015). It remains to be investigated patterns of activation in the mentalizing system during an eye gaze task in those with psychotic BD. Better understanding of the neural underpinnings of altered social cognition would inform the design of targeted interventions to improve functioning.

## 2. METHODS

### 2.1 Participants

Data of 14 individuals with DSM-IV-TR bipolar disorder with psychotic features (BD) and 21 demographically matched healthy controls (HC) were included in this report. The study was conducted in accordance with the protocol approved by the Institutional Review Board of the University of Michigan Medical School. Written informed consent was obtained from each participant after full explanation of the study was provided. Details of participants’ demographic and clinical characteristics can be found in Online Supplemental Information.

### 2.2 Eye Gaze Perception Task

Participants performed the Eye Gaze Perception Task during fMRI. They viewed faces with different gaze angles. For each face, they pressed a button to indicate perceived self-directed gaze (yes or no) or the gender of the face (male or female). Details of the task, along with behavioral data analysis and results, are described in the Online Supplemental Information.

### 2.3 Data Acquisition & Preprocessing

MRI scanning occurred on a 3T GE MR 750 Discovery scanner. A T1-weighted image was acquired in the same prescription as the functional images. Functional images were acquired with a T2*-weighted, reverse spiral acquisition sequence (gradient echo, TR=2000 ms, TE=30 ms, FA=90 degrees, FOV=22 cm, 40 slice, 3mm thick/0mm skip, equivalent to 64 × 64 voxel grid). Subsequently, a high resolution T1 scan (AxF SPGR, FOV=26 cm, TI=500 ms, FA=15 degree, BW=31.25, 256 × 256 matrix, 128 slices, 1.2 mm interleaved with no skip) was obtained for anatomic normalization.

fMRI data were processed using typical methods in Statistical Parametric Mapping, including slice time correction, realignment to the 10th volume acquired, co-registration of time series of functional volumes with high resolution T1 image, spatial normalization to the MNI152 brain, followed by spatial smoothing with an 8 mm isotropic Gaussian kernel. See Supplemental Information for additional details.

### 2.4 fMRI Data Modeling and Identification of Regions of Interest (ROIs)

The anatomically normalized time series was regressed on 2 regressors of interest (Gaze trials, Gender trials) and 27 nuisance regressors, convolved with a canonical hemodynamic response function. Beta values (first eigenvector of a 10 mm radius sphere) from the Gaze-Gender contrast were then extracted from three regions of interest (ROIs)—the medial prefrontal cortex (mPFC) and the bilateral temporo-parietal junctions (TPJs)—based on the coordinates derived from a meta-analysis of fMRI studies of the mentalizing brain system (Van Overwalle, 2009; Van Overwalle and Baetens, 2009).

### 2.5 Statistical Analysis

Differential brain activation in the three ROIs between HC and BD was examined using independent-samples t-tests. To evaluate the functional relevance of brain activation in these three ROIs, canonical correlation analysis (CCA) was conducted to delineate the collective relationship between preferential brain activation for gaze discrimination (vs. gender identification) in these ROIs and two functional measures (neurocognition and social cognition).

## 3. RESULTS

Behavioral results are summarized in Supplemental Results; in short, no significant differences were observed between HC and BD. The fMRI results are shown in Figure 1. While HC showed similar or more activation during gaze (vs. gender) trials in the three ROIs, BD showed significantly reduced preferential activation for gaze discrimination in mPFC and left TPJ.

CCA revealed that the relationship between preferential brain activation for gaze discrimination (vs. gender identification) and functional measures was statistically significant, Wilk’s Λ = 0.645, *p* = .035, yielding one significant canonical function such that more preferential brain activation for gaze discrimination across the three ROIs were collectively associated with better neurocognitive and social cognitive functioning (Supplemental Information Table S2).

**Figure 1.**
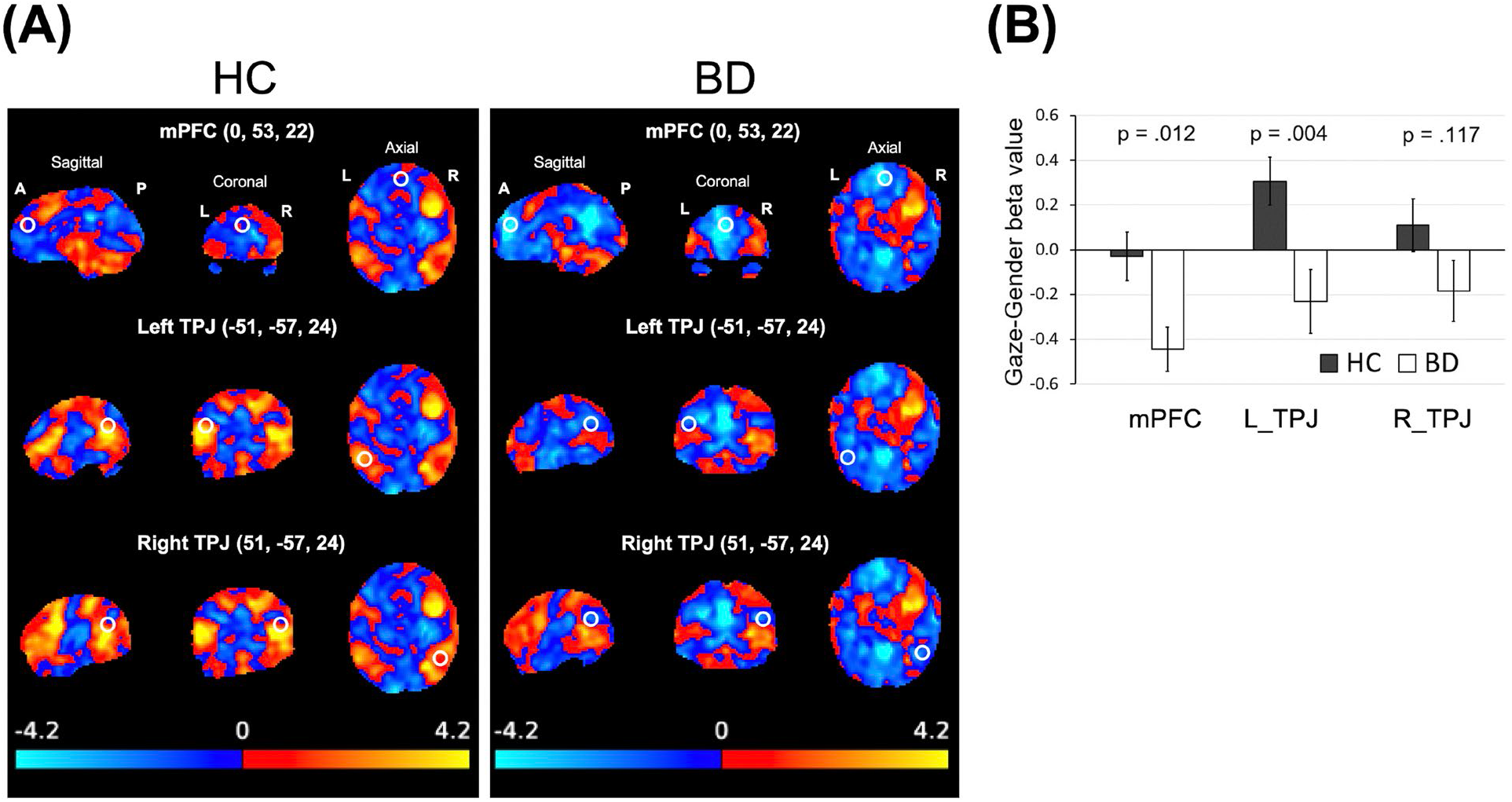
Brain activation during gaze discrimination vs. gender identification. (A) Whole-brain activation map (unthresholded) of the Gaze-Gender contrast for healthy control (HC; left panel) and bipolar (BD; right panel) participants. The white circles indicate the regions of interest. (B) Extracted beta values of the Gaze-Gender contrast. mPFC = medial prefrontal cortex; L_TPJ = left temporo-parietal junction; R_TPJ = right temporo-parietal junction.

## 4. DISCUSSION

This study aimed to examine the neural correlates of altered eye gaze processing in BD with psychosis. Specifically, task-related activation in three core brain regions constituting the mentalizing brain system—bilateral TPJs and mPFC—were compared between individuals with BD and healthy comparison participants. Similar to prior findings on other mentalizing tasks (Malhi et al., 2008; Willert et al., 2015), those with BD showed reduced preferential activation in mPFC and left TPJ during gaze discrimination, suggesting dysfunctional recruitment of brain regions supporting this social cognitive process. Given the putative functions of these brain regions to support complex social cognitive processes, the results are also consistent with prior behavioral findings suggesting the role of self-referential bias in social dysfunction associated with BD (Yao et al., 2018). Moreover, reduced activation was associated with poorer neurocognitive and social cognitive functioning. Together these preliminary results suggest that diminished activation in the mentalizing brain system when processing social cues may be a marker of poorer functional outcome in BD.

Larger sample sizes are needed to replicate and confirm these findings. Further, determining whether the aberrant activation observed in this study is related to psychosis specifically, or is common among non-psychotic affective disorders as well, is a logical next step. The null mPFC activation value in HC may be because the gaze task recruits more posterior mPFC than the meta-analysis comprising diverse mentalizing tasks, which were more likely to activate dorsal mPFC. Finally, further study is warranted to investigate whether modulating these neural targets using interventions such as cognitive therapy, cognitive training, and/or brain stimulation may improve social functioning among individuals with BD.

## Data Availability

The data for this study are not publicly available because permission was not obtained from participants for open-access data sharing.

## Acknowledgment

This research was supported by the National Institutes of Health (5KL2TR000434 to I.F.T.), the National Institute of Mental Health (5K23MH108823 and R01MH122491 to I.F.T.; R01MH118634 to S.F.T.), and the Heinz C. Prechter Bipolar Research Fund (to M.G.M.).

## Contributors

**Ivy Tso:** Conceptualization, methodology, funding acquisition, project administration, investigation, data curation, formal analysis, visualization, supervision, writing – original draft; **Cynthia Burton:** Investigation, writing – original draft; **Carly Lasagna:** Methodology, visualization, writing – review & editing; **Saige Rutherford:** Software, formal analysis, visualization, writing – review & editing; **Beier Yao:** Investigation, data curation, writing – review & editing; **Scott Peltier:** Methodology, investigation, writing – review & editing; **Timothy Johnson:** Methodology, supervision, writing – review & editing. **Melvin McInnis:** Project administration, resources, writing – review & editing; **Stephan Taylor:** Methodology, funding acquisition, project administration, supervision, writing – review & editing. All authors provided feedback on the final version of the manuscript.

