## Supplemental Information for "Aberrant Activation of the Mentalizing Brain System During Eye Gaze Discrimination in Bipolar Disorder"

#### Online Supplemental Information

### SUPPLEMENTAL METHODS

**Participants.** Sixteen individuals with DSM-IV-TR bipolar disorder with psychotic features (BD) and 22 healthy controls (HC) completed the study. Participants were recruited through community advertisements and referrals by researchers from other research programs (McInnis et al., 2018) or clinicians of local mental health clinics. Diagnoses were established using the Structured Clinical Interview for DSM-IV Axis-I Disorders (SCID-I) (First et al., 1995). Inclusion/exclusion criteria were: age 18 to 55 years old; capable of giving informed consent; normal or corrected-to-normal vision; no significant medical or neurological illness; no history of closed head injury with neurological sequella; pregnancy; no contraindications to MRI (e.g., metal objects in body); and no alcohol/substance abuse in the past month or dependence in the past 6 months. Additional exclusion criteria for HC included: past and current Axis I disorder; history of psychotic or bipolar disorder among first-degree relatives; and current Beck Depression Inventory (BDI-II) (Beck and Steer, 1993) score > 8.

Data of three participants were excluded from the analyses for not responding to most trials (one BD), having excessive motion in 2 out of 3 runs of the fMRI task (one BD), and excessive scanner artifacts (one HC). The remaining 14 BD and 21 HC participants were well matched demographically (see Table S1 below).

**Assessments.** Participants completed the BDI-II and Altman Self-Rating Mania Scale (ASRM) (Altman et al., 1997) as measures of depression and mania symptoms, respectively. Neurocognition was assessed using the MATRICS Consensus Cognition Battery (MCCB) (Nuechterlein et al., 2008). Social cognition was assessed with the Mayer-Salovey-Caruso Emotional Intelligence Test (MSCEIT) (Mayer et al., 1999).

**Eye Contact Perception Task (GAZE).** Stimuli were forward-facing faces with 9 varying gaze directions presented in pseudo-randomized order. These gaze angles represent levels of eye-contact signal strength, from 0.2, 0.3, …, to 1.0 for averted to direct gaze. For each face, participants indicated perceived eye contact (Gaze trials: "Looking at you? Yes/No") or gender (Gender trials: "Gender? Male/Female") by pressing a button. Sample stimuli and design specifics are illustrated in Figure S1.

**fMRI data acquisition.** A T1-weighted image was acquired in the same prescription as the functional images to facilitate co-registration. Functional images were acquired with a T2*-weighted, reverse spiral acquisition sequence with excellent signal recovery in areas of high susceptibility artifact (gradient echo, TR=2000 ms, TE=30 ms, FA=90 degrees, FOV=22 cm, 40 slice, 3mm thick/0mm skip, equivalent to 64 × 64 voxel grid). Participants underwent 3 runs, with each run containing 12 task blocks and 11 fixation blocks. Each run had 228 volumes plus 4 initial, discarded volumes to allow for equilibration of scanner signal, with isotropic voxels 3 mm on edge. After acquisition of functional volumes, a high resolution T1 scan (AxF SPGR, FOV=26 cm, TI=500 ms, FA=15 degree, BW=31.25, 256 × 256 matrix, 128 slices, 1.2 mm interleaved with no skip) was obtained for anatomic normalization.

**fMRI data preprocessing.** fMRI data were processed using typical methods in Statistical Parametric Mapping (SPM12, Wellcome Institute of Cognitive Neurology, London). Slice time was done using sinc-interpolation, weighted by a Hanning kernel in time. Then all scans were realigned to the 10th volume acquired during each scan. Scans with movement exceeding either 1 voxel or 2-degree rotation within a run were discarded. There were no significant differences in the average framewise displacement between groups (HC = 0.14 ± 0.08 mm; 0.15 ± 0.084 mm; $t$ = -0.47, $p$ =0.63). Time series of functional volumes were then co-registered with high resolution T1 image, spatially normalized to the MNI152 brain, and then spatially smoothed with an 8 mm isotropic Gaussian kernel.

**fMRI Data Modeling.** First-level analysis began with applying a high pass filter (128 s) to the anatomically normalized time series and then regressing time series on regressors convolved with a canonical hemodynamic response function. The regressors included 2 regressors of interest (Gaze trials, Gender trials) and 27 nuisance regressors including 3 runs and 24 motion parameters (6 for each translation/rotation direction, their quadratic terms, and first and second derivatives).

**Gender identification accuracy and reaction time.** Accuracy in gender identification, as a proxy of attention during the task, was compared between BD and HC with $t$-test. Reaction time (RT) for Gender and Gaze trials was examined with separate linear mixed models (LMMs). The models included fixed effects of: Group, Eye-Contact Signal Strength ("Signal Strength" from now on), Group × Signal Strength, Signal Strength$^2$, and Group × Signal Strength$^2$; the quadratic terms were to address the non-linear effect of signal strength on RT. Intercept was modeled as a random effect, and Signal Strength was treated as a repeated-measures and continuous variable.

**Derivation of psychophysical gaze perception metrics.** We derived two metrics of gaze perception for each participant based on their behavioral responses on the gaze perception task: slope (which indexes perceptual precision) and threshold (which indexes self-referential bias). See Figure S2 for an illustration. For each participant, the number of eye-contact endorsement ("yes" responses), $Y_{ij}$, for each eye-contact signal strength was modeled as a random variable that follows a binomial probability distribution dependent on $\theta_{ij}$ (an unknown, true value between 0 and 1) and the number of completed trials, $T_{ij}$:

$$Y_{ij} \sim Binomial(\theta_{ij}, T_{ij})$$

*where i = participant index,*

*j = index corresponding to the 9 eye-contact signal strengths*

As demonstrated in our prior work (Tso et al., 2012), the probability of perceiving eye contact varies with eye-contact signal strength and approximates a logistic function. Therefore, $\theta_{ij}$ was linked to eye-contact signal strength, $X_j$, via a logit function using two free parameters, $\alpha_i$ and $\beta_i$:

$$\text{logit}(\theta_{i,j}) = \alpha_i + \beta_i \cdot X_j$$

Parameters $\alpha_i$ and $\beta_i$ were then modeled to come from a normal distribution centered around the mean of the participant's corresponding group $k$:

$$\alpha_i \sim Normal(\mu_{\alpha k}, \tau_{\alpha k})$$

$$\beta_i \sim Normal(\mu_{\beta k}, \tau_{\beta k})$$

*where k = 1 (HC) for i = 1 – 21*

*k = 2 (BD) for i = 22 – 35*

In the Bayesian estimation, uninformative priors were assigned to the parameters making up of the group-level normal distributions. Specifically, flat priors were used for the mean parameters and gamma(1,1) priors were used for the precision parameters. Individual ($\alpha_i$, $\beta_i$) and group parameters ($\mu_{\alpha k}$, $\mu_{\beta k}$, $\tau_{\alpha k}$, $\tau_{\beta k}$) were used to compute the two gaze perception metrics of interest for each individual (threshold = - $\alpha_i/\beta_i$; slope = $\beta_i/4$) and each group (threshold = -$\mu_{\alpha k}/\mu_{\beta k}$; slope = $\mu_{\beta k}/4$). The joint posterior distribution function of the parameters was estimated using the Gibbs sampling algorithm (Geman and Geman, 1984) as implemented in WinBUGS (Version 1.4) (Spiegelhalter et al., 2003). Two Markov chains of 100,000 samples (with the first 4,000 discarded as burn-in) were generated to approximate the posterior probability density distributions of the parameters.

#### SUPPLEMENTAL RESULTS

**Gender identification accuracy and reaction time (RT).** BD (0.84 ± 0.07) and HC (0.84 ± 0.10) showed similar gender identification accuracy, $t(33) = 0.04$, $p = .97$. Results of RT are illustrated in Figure S3. For Gaze trials, both BD and HC needed more time to respond to more direct gaze and the longest to respond to gaze in the "ambiguous" zone (middle area of the gaze continuum), but they were not different in overall RT. As expected, RT for Gender trials showed no relationship with Signal Strength across the two groups, but BD were slower in responding in general.

**Gaze perception metrics.** The threshold and slope of the two groups were not credibly different, such that the posterior probability of higher threshold in HC relative to BD was 69.2% for threshold and that of steeper slope in HC than BD was 69.9%. See Figure S4.

McInnis, M.G., Assari, S., Kamali, M., Ryan, K., Langenecker, S.A., Saunders, E.F.H., Versha,

K., Evans, S., O'Shea, K.S., Provost, E.M., Marshall, D., Forger, D., Deldin, P., Zoellner, S., Harrington, G.J., Bame, M., Bertram, H., Brucksch, C., Chun, J., Cochran, A., DeLong, C., Easter, R., Ellingrod, V., Foster, V., Kang, G.E., Ghaziuddin, N., Gideon, J., Greden, J., Grimm, C., Gross, M., Jenkins, P., Kelly, M., Khorram, S., Martinez, E., Mueller, S., O'Donnell, L., Preiser, B., Prossin, A., Thompson, S.B., Williams, A., 2018. Cohort profile: The Heinz C. Prechter longitudinal study of bipolar disorder. Int. J. Epidemiol. 47, 28-28n. https://doi.org/10.1093/ije/dyx229

Nuechterlein, K.H., Green, M.F., Kern, R.S., Baade, L.E., Barch, D.M., Cohen, J.D., Essock, S., Fenton, W.S., Frese, F.J., Gold, J.M., Goldberg, T., Heaton, R.K., Keefe, R.S.E., Kraemer, H., Mesholam-Gately, R., Seidman, L.J., Stover, E., Weinberger, D.R., Young, A.S., Zalcman, S., Marder, S.R., 2008. The MATRICS Consensus Cognitive Battery, Part 1: Test Selection, Reliability, and Validity. Am. J. Psychiatry 165, 203–213. https://doi.org/10.1176/appi.ajp.2007.07010042

Spiegelhalter, D.J., Thomas, A., Best, N., Lunn, D., 2003. WinBUGS User Manual Version 1.4 [WWW Document]. URL https://faculty.washington.edu/jmiyamot/p548/spiegelhalter winbugs user manual.pdf

Tso, I.F., Mui, M.L., Taylor, S.F., Deldin, P.J., 2012. Eye-contact perception in schizophrenia: Relationship with symptoms and socioemotional functioning. J. Abnorm. Psychol. 121, 616–627. https://doi.org/10.1037/a0026596

Table S1. Participant Characteristics

| | BD ($n$ = 14) Mean ± SD | HC ($n$ = 21) Mean ± SD | t / $\chi^2$ | $p$ |
|---|---|---|---|---|
| *Demographics* | | | | |
| Age | 32.1 ± 8.9 | 31.0 ± 12.5 | -.307 | .761 |
| Sex (male/female) | 6 / 8 | 11 / 10 | .305 | .581 |
| Education, years | 16.4 ± 2.1 | 15.6 ± 1.9 | -1.268 | .214 |
| Parental education, years | 17.0 ± 2.1 | 15.7 ± 3.7 | -1.189 | .243 |
| *Clinical Assessments* | | | | |
| BDI | 7.9 ± 10.3 | -- | -- | -- |
| ASRM | 3.0 ± 2.6 | -- | -- | -- |
| *Functional Assessments* | | | | |
| MCCB | 49.9 ± 29.4 | 71.7 ± 23.3 | 2.445 | .020 |
| MSCEIT | 103.6 ± 13.8 | 115.0 ± 19.3 | 2.027 | .051 |

Note. BDI = Beck Depression Inventory; ASRM = Altman Self-rating Mania Scale; MCCB = MATRICS Consensus Cognitive Battery; MSCEIT = Mayer-Salovey-Caruso Emotional Intelligence Test.

Table S2

*Canonical solution for preferential brain activation for gaze discrimination predicting functioning*

| Variable | Canonical Function | | |
|---|---|---|---|
| | Coeff | $r_s$ | $r_s^2$ |
| Preferential activation for gaze discrimination | | | |
| mPFC | 0.433 | .830 | 68.89% |
| Left TPJ | 0.827 | .940 | 88.36% |
| Right TPJ | -0.230 | .593 | 35.16% |
| Functioning | | | |
| Neurocognition (MCCB) | 0.484 | .791 | 62.57% |
| Social cognition (MSCEIT) | 0.684 | .902 | 81.36% |

*Note.* Coeff = standardized canonical function coefficient; $r_s$ = structure coefficient; $r_s^2$ = structure variance explained.

a)

Face stimuli removed per medRxiv policy.
Please contact the corresponding author for access to stimuli.

b)

Figure of task design removed per medRxiv policy (because it contained sample images of face stimuli).
Please contact the corresponding author for access to details of the task.

**Figure S1. Eye Gaze Perception Task. a)** Sample face stimuli with 9 varying gaze angles. **b)** fMRI design: Gaze discrimination and gender identification tasks were presented in blocks (19.8 – 24.4 s), alternating with a rest block (11.4 – 15.6 s) in between. Each face was presented for 1.5 s, and the 6 faces within each block were separated by a random jitter (1.6 – 3.9 s). Totally 108 trials, divided into 3 runs, were presented for each task (Gaze, Gender).

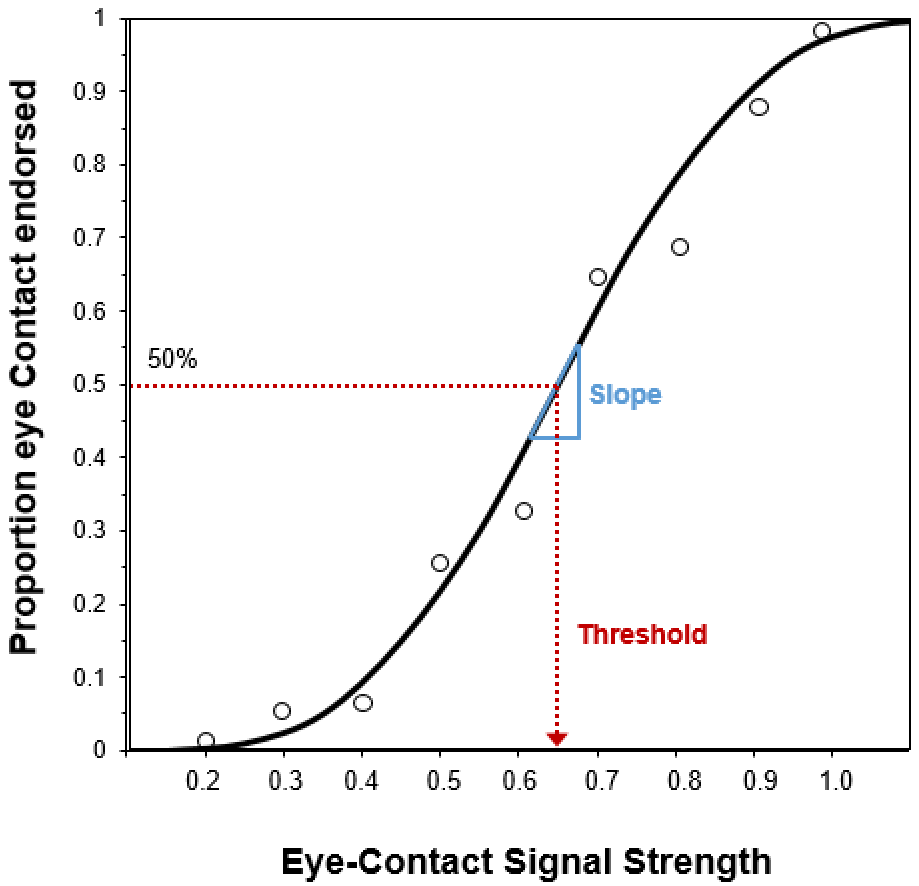


**Figure S2. Probability of self-directed gaze endorsement as a logistic function of eye-contact signal strength.** Round markers represent observed data (proportion of the number of trials in which self-directed gaze was endorsed out of the total number of trials completed at each signal strength). The curve represents a theoretical, logistic function generative of the data. The threshold (*x* value at *y* = 50%) was used to index self-referential tendency, and the slope of the curve at *y* = 50% was used to index visual perceptual sensitivity.

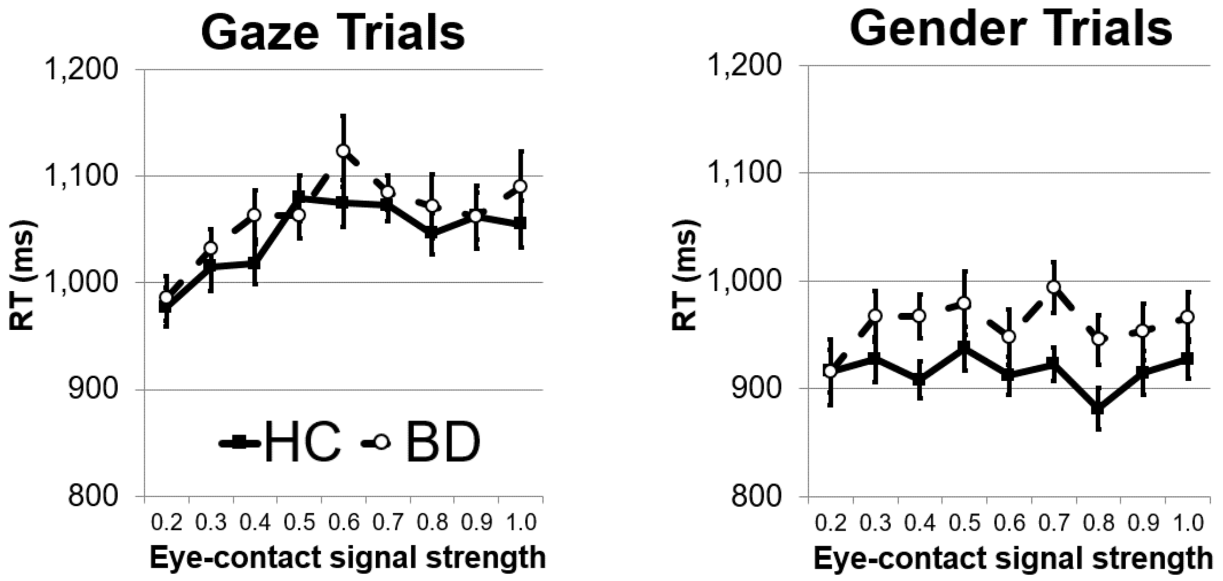


**Figure S3. Reaction time (RT) in the gaze processing task among healthy control (HC) and bipolar disorder (BD) participants.** Left panel: RT for Gaze trials displayed a linear, $F(1, 277.5) = 32.26$, $p < .001$, as well as a quadratic effect of Signal Strength, $F(1, 277.5) = 25.31$, $p < .001$. These indicate that RT tended to be longer for gaze with stronger eye-contact signal, but the longest for ambiguous gaze. HC and BD were not significantly different in overall RT for Gaze trials, $F(1, 44.2) = 20.87$, $p = .36$. The linear or quadratic effects of Signal Strength also did not differ between HC and BD, Signal Strength × Group: $F(1, 277.5) = 0.07$, $p = .79$; Signal Strength $^2$ × Group: $F(1, 277.5) = 0.05$, $p = .82$. Right panel: RT for Gender trials was overall longer for BD than HC, $F(1, 39.90) = 4.42$, $p = .042$. As expected, there were no linear, $F(1, 277.1) = 0.20$, $p = .66$, nor quadratic relationship with Signal Strength, $F(1, 277.1) = 2.18$, $p = .14$. There was also no Signal Strength × Group interaction, $F(1, 277.1) = 1.79$, $p = .18$, nor Signal Strength $^2$ × Group interaction, $F(1, 277.1) = 3.31$, $p = .07$. Note: for eye-contact signal strength, larger numbers indicate more direct eye gaze.

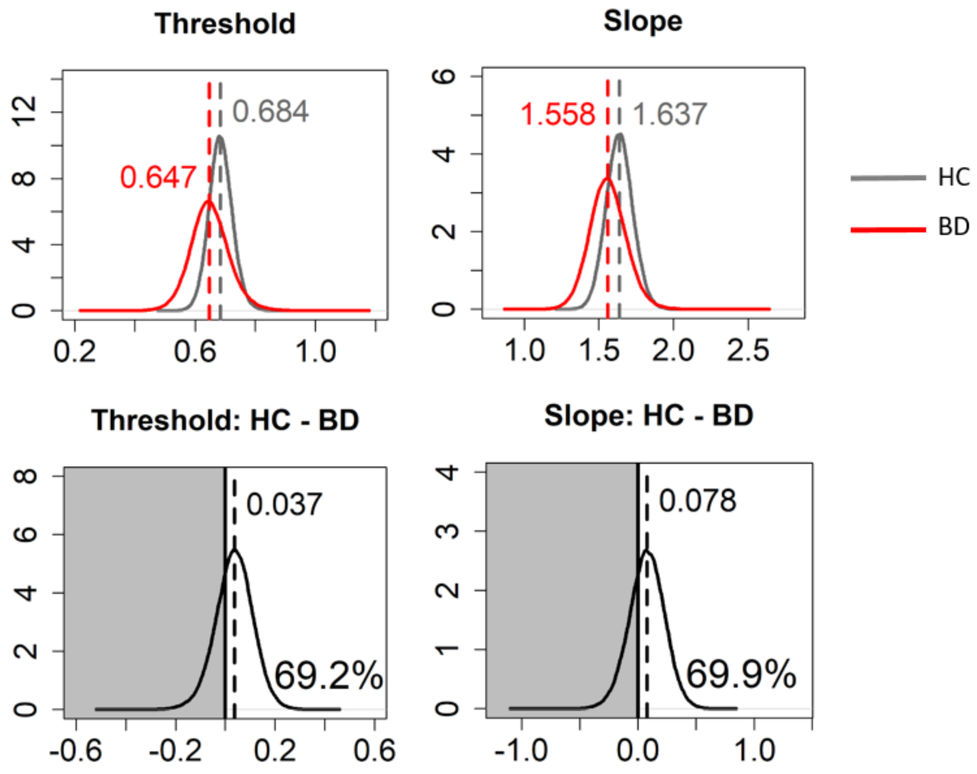


**Figure S4. Posterior probability density plots of the gaze perception metrics in healthy controls (HC) and bipolar disorder (BD).** Dashed vertical lines and numbers at the top indicate median values of Markov Chain Monte Carlo (MCMC) samples. Numbers at the bottom right (bottom panel) indicate the posterior probability of the HC > BD group difference (i.e., area under the curve in the white area).
